# Tau Progression Index (TPI): An individualized, clinically applicable, multimodally-derived score to predict AD progression

**DOI:** 10.1101/2025.01.14.25320550

**Authors:** Seyed Hani Hojjati, Gloria C. Chiang, Tracy A. Butler, Bardiya G. Yazdi, Sindy Ozoria, Peter Chernek, Jenseric Calimag, Yaakov Stern, Devangere P. Devanand, Sonja Blum, Silky Pahlajani, Christian G. Habeck, Hengda He, Nancy Foldi, Mohammad Khalafi, Qolamreza R. Razlighi

## Abstract

While amyloid-beta (Aβ) and tau are known to be the key neuropathological hallmarks of Alzheimer’s disease (AD), imaging and postmortem studies highlight significant variability in disease severity and progression even in subjects with similar burdens of Aβ and tau accumulation. This uncertainty in the trajectory of disease progression results in substantial challenges for patients and their families in understanding disease prognosis, as well as the research community in understanding heterogeneity in the rate of Aβ and tau accumulation, cognitive decline, and likelihood of progressing to dementia. In this study, we developed and validated the Tau Progression Index (TPI); a multimodally derived score that leverages strongly supported hypotheses and models of tau spread, to serve as a reliable, and clinically applicable biomarker of AD progression. TPI has 4 components: 1) Remote interaction (TPI-ri) interactions between Aβ and tau pathologies in regions that spatially separate; 2) Local interaction (TPI-li) between co-localized Aβ and tau pathology; 3) Functional connectivity (TPI- fc) is based on functional magnetic resonance imaging (fMRI)-measured inter-regional functional connections facilitate the spread of tau in the brain; and 4) Structural connectivity (TPI-sc) based on diffusion MRI (dMRI)-measured inter-regional structural connectivity. Importantly, all prior models of tau spread in AD were based on group-averaged structural and functional connectivity, derived from healthy and often young subjects. However, brain connectivity patterns are extremely subject-specific, often referred to as a human connectome fingerprint, and are significantly altered by aging and neurodegenerative diseases. Therefore, we have shown that TPI computed by subject-specific connectomes is crucial to accurately model subject-specific trajectories of tau spread.

## 1. Introduction

Alzheimer’s disease (AD) is the most common neurodegenerative disease, currently affecting over 6 million adults in the US and projected to affect nearly 13 million Americans by 2050^1^. AD is a dual proteinopathy such as amyloid-beta (Aβ) plaques and intracellular neurofibrillary tau tangles,^2–4^ with evidence showing that tau accumulation is more associated with neurodegeneration and cognitive decline than Aβ.^5–8^ However, multiple post-mortem^9,10^ and imaging^11,12^ studies have shown that both Aβ and tau can be present in up to 30% of cognitively normal individuals. Furthermore, prognosis is variable even with a similar burden of global tau deposition in the brain, since the likelihood of converting from cognitively normal (CN) to mild cognitive impairment (MCI) within two years has a wide range of 33 to 83%^13^. Because of these differences in the rates of progression and patterns of tau spread among individuals with AD pathology, it is extremely challenging to develop targeted therapeutic interventions and counsel families on disease prognosis.

Recent literature has reported that the human brain connectomes can be used to predict the spread of tau across regions^14–19^, based on the concept of transneuronal spread of tau^15,18,20^. For instance, Frontzkowski et al., 2022^14^ introduced the “*tau hub ratio*” where tau deposition in regions with maximal number of functionally connected regions to the rest of the brain “regional hubs” tend to spread more than the tau in non-hub regions. Similarly, Jin Lee et al., 2022^21^ showed that structural connectivity could facilitate remote interactions between Aβ and tau in connected regions, contributing to the spread of tau to the rest of the brain. However, all prior studies, to our knowledge, used group-average connectomes from young, healthy cohorts. Because connectomes are significantly altered by aging^22–26^ and neurodegenerative diseases^27–30^, and are highly individualized across subjects^31^, a subject-specific connectome is crucial to accurately model heterogeneous trajectories of tau spread. The recent development of multi-band acquisition^32,33^ and sophisticated MR processing and analysis techniques by us^34–36^ and others^37–41^ now allow reliable extraction and definition of functional and structural connectomes unique to an individual.

In this study, we propose to develop a tau progression index (TPI), which allows us to accurately predict a subject-specific trajectory of disease progression that can be targeted by therapies, monitored in clinical trials and used to provide a more specific prognosis. Importantly, all prior models of tau spread in AD were based on group-averaged structural and functional connectivity, derived from healthy and often young subjects^14,16,19,21^. TPI has 4 components: 1) Remote interaction (TPI-ri) is based on work by us^42,43^ and others^21,44–46^ that inter-regional interactions between Aβ and tau pathologies in regions that are functionally and/or structurally connected, but spatially separate, strongly promote AD onset and progression; 2) Local interaction (TPI-li) is based on our demonstration using AD neuroimaging initiative (ADNI) data that spatially co-localized Aβ and tau pathology strongly predicted conversion from HC to MCI as well as MCI to AD^47^; 3) Functional connectivity (TPI-fc) is based on extensive work by us^48,49^ and others^14–16,50^ showing that functional magnetic resonance imaging (fMRI)-measured inter-regional functional connections facilitate the spread of tau in the brain, with tau in regions with the highest number of functionally connected regions spreading more rapidly and extensively than tau in less connected regions; and 4) Structural connectivity (TPI-sc) is based on works highlighting a robust contribution of diffusion MRI (dMRI)-measured inter-regional structural connectivity to tau progression^15–18,21^.

We aim to use TPI to detect individuals with a higher risk of tau elevation. Using the least absolute shrinkage and selection operator (LASSO), our final TPI (TPI_final_) differentially weights the unique and shared contribution of each TPI component in predicting future tau spread. By doing so, our TPI would provide a step towards guiding personalized treatment, by incorporating spatial patterns of tau deposition and brain connectomes into a prognostic tool for patient counseling and clinical trials.

## 2. Methods

### 2.1. Participants

We have recruited exclusively from ongoing studies that are already acquiring Aβ and tau PET scans with harmonized protocols across New York-Presbyterian/Columbia and Cornell medical centers. There are more than 650 participants across our studies who have already been scanned with both PET tracers (^18^F-Florbetaben, ^18^F-MK4260) as well as the required magnetic resonance imaging (MRI) scans. From this current pool of 650+ participants, currently, 112 individuals met our neuroimaging inclusion criteria (See next section). From 112 identified participants, 27 individuals (age 68.25 ± 6.02 years; 15 females; 10 MCI and 17 CN) have already been followed for 2∼3 years which was used in this study to evaluate our proposed TPI measurement. In addition, all individuals underwent MRI, and functional and diffusion (MRI) evaluations covering medical and neuropsychological assessments. All study procedures are approved by the local institutional review board in Columbia and Cornell University medical centers. All participants signed the informed consent form before participating in the study and they were compensated for the time they spent on the study.

Another cohort of young participants (20 ∼ 40 years) were recruited in Weill Cornell medicine using the market mailing method and scanned with the same protocols as above. This cohort was exclusively used for defining our normative samples to identify Aβ and Tau positivity.

### 2.2. Primary Inclusion/Exclusion Criteria

To optimally evaluate our proposed TPI measurement, participants with early deposition of the A<u>β</u> and/or tau are required. This is because, if the tau is already spread to most of the brain regions in a participant, then that individual might not be suitable for evaluation of our proposed TPI for predicting future tau accumulation. Inversely, if a participant has no tau accumulation in the baseline and follow-up, natural that participants be useful in evaluating our proposed TPI. Therefore, we have imposed both lower and higher thresholds of tau spread in our inclusion criteria as follows: 1) *<u>Tau burden:</u>* Subjects must have greater than 5 but less than 50 tau-positive (tau+) regions (out of 216 regions; 200 Schaefer atlas + 16 subcortical regions). We defined a tau+ region to be MK6240 SUVR_reg_>95^th^ percentile of our cognitively healthy, young (20-40 years of age) cohort’s regional SUVR distribution. 2) *<u>A</u>*<u>β</u> *<u>deposition:</u>* The minimum required amount of Aβ deposition is defined as SUVR_reg_>95^th^ percentile of our young, healthy cohort’s regional SUVR distribution in more than 10 regions.

We have successfully used this novel approach to define our regional cut-off threshold using data from a young cohort in our recent publication^42,43^. 3) *<u>Clinical/cognitive status:</u>* We enrolled subjects who are either cognitively normal (preclinical AD) or have MCI. We exclude subjects with dementia at baseline because this would make it more difficult to measure progression/change, which is the focus of this project. Subjects for this study must have a CDR global score of 0 (cognitively normal) or 0.5 (MCI in accordance with NIA criteria^51^) based on an interview/examination with a cognitive neurologist.

### 2.3. Neuroimaging acquisitions

All PET scans are acquired on three different Siemens Biograph mCT–S scanners in dynamic and 3D imaging mode. The tau PET and Aβ PET scans took place over 2 different sessions. Longitudinal scans are always acquired on the same scan as the baseline. Pre-injection and post-injection vital signs are collected and recorded.

#### Aβ Imaging

All participants at the baseline visit had ^18^F-Florbetaben PET scans to assess Aβ burden. Participant preparation consists of intravenous catheterization followed by the injection of 8.1mCi ± 20% (300 Mbq) of the tracer administered as a slow single intravenous bolus at 60 sec or less (6sec/mL max). The 3D imaging begins 90 minutes after injection of the tracer. A low-dose CT scan for attenuation correction of the PET data has been acquired. Brain images were acquired in 4 x 5-minute frames over a period of 20 minutes.

#### Tau Imaging

All participants at baseline and follow-up visits had ^18^F-MK6240 PET scans to assess tau deposition. Participant preparation consists of intravenous catheterization or butterfly for injection. Injection of 185 MBq (5 mCi) ± 20%, (maximum volume 10 mL) administered as a slow single intravenous bolus at 60 sec or less (6sec/mL max). A low-dose CT scan for attenuation correction of the PET data acquired. The 3D imaging begins 90 minutes after tracer injection. Brain images were acquired in 6 x 5-minute frames over a period of 30 minutes.

#### MRI acquisition parameters

Both baseline and follow-up MR images were acquired in the same research-dedicated Siemens 3.0 Tesla Prisma scanner with a 64-channel head-coil and the same pulse sequences and acquisition protocols. Quarterly calibration with Magphan S162 (https://www.phantomlab.com/magphan-s162) ensures consistent, high quality at baseline and follow-up acquisitions. Each MRI session for baseline and follow-up started with a high resolution (0.5mm^3^) MPRAGE structural scan [TR/TE = 2400/2.96 ms, flip angle = 9°; FOV = 25.6×25.6 cm, matrix size = 256×256, and 208 sagittal slices]. Then, 2 resting-state fMRI scans [TR/TE = 1008/37 ms; flip angle = 52°; FOV = 20.8×20.8 cm; matrix size = 104×104; voxel size = 2x2x2 mm; 72 axial slices, and MB=6] each lasting 10 minutes have been performed with opposite phase encoding directions. Participants were instructed to lie still with their eyes open, stay awake, and clear their minds. Finally, 2 dMRI scans [TR/TE=3230/89.20 ms; flip angle = 78°; FOV=21x21 cm; matrix size = 140x140; voxel size = 1.5x1.5x1.5 mm, 92 axial slices; 2 b-values = 0, and 3000 s/mm^2^, and MB=6] (5 min, 37 sec each) with 98 directions were acquired with opposite phase encoding directions.

### 2.4. Neuroimage data processing

#### Structural Image Processing

The T1-weighted MPRAGE scans have been processed using FreeSurfer (http://surfer.nmr.mgh.harvard.edu/), an automated segmentation and cortical parcellation software package^52,53^. All participants’ cortical surfaces were visually inspected/corrected by a demographic-blind technician. In the case of discrepancy, manual editing of the white and gray matter borders was conducted per the FreeSurfer manual editing guidelines. Freesurfer segments the cortex into 33 different gyral-/sulci-based regions in each hemisphere according to the Desikan-Killiany atlas^54^, and calculates the cortical thickness at each vertex, which is at 1x1 mm resolution. The maps produced can detect submillimeter differences between groups^55^. In addition, the sub-cortical regions are also segmented to give 37 subcortical regional masks. Intracranial brain volume (ICV) is measured using BrainWash (an automatic multi-atlas skull-striping software package)^56^.

We processed the longitudinal T1-weighted MPRAGE images using the FreeSurfer longitudinal pipeline, which is implemented to detect small or subtle changes over time. This processing pipeline is robust to the initialization points which often generate small variations in the results of the optimization processes.^57^

Spatial normalization of each participant’s FreeSurfer space scans to the MNI template is done using ANTS non-linear registration tools using 100x100x100 iteration.

#### Quantification Process for PET Data

An in-house developed fully automatic quantification method has been implemented and validated with histopathological data^58^ and used in numerous published studies for quantification of PET scans^59–62^. Briefly, it starts by aligning dynamic PET frames (6 frames in tau-PET and 4 frames in Aβ-PET) to the first frame using rigid-body registration and averaging them to generate a static PET image. Each individual’s structural T1 image in FreeSurfer space is also registered to the same participant’s static image using normalized mutual information and 6 degrees of freedom to obtain a rigid-body transformation matrix to transfer all FreeSurfer regional masks to static PET image space. These regional masks in static PET space are used to extract the regional PET data. The standardized uptake value (SUV) is calculated at selected regions and then normalized to cerebellum gray matter to derive the standardized uptake value ratio (SUVR). In the volumetric analysis of the Florbetaben scan, the spill-in signal from nonspecific binding in white-matter is attenuated by discarding the uptake in the gray-matter voxels located immediately adjacent to the white-matter volume, both in the cerebellum for computing the reference region uptake and in the cerebral cortex for obtaining cortical regions’ SUVR. For MK6240, the non-specific binding is mainly in the meninges, so the same approach is used to discard all voxels on the convex hull of the brain surface. Using the ANTS worping field and the SUVR scan in FreeSurfer space all regional uptake for Schaefer atlas is also computed.

#### Processing fMRI Data

Our fMRI processing pipeline was a combination of steps from the FSL toolbox and in-house-developed techniques. Briefly, in-house developed slice timing correction is applied to the raw fMRI time series to account for the difference in the acquisition delay between slices^35,63,64^. At the same time, motion parameters were estimated on raw fMRI scans using rigid-body registrations performed on all the volumes in reference to the first volume. A k-space-based motion correction technique^34^ was applied. Additionally, the first volume was extracted from the two fMRI scans with opposite phase encoding directions used to estimate the geometric distortion correction field using a susceptibility-induced distortions correction technique called TOPUP provided in the FSL software package^65^. Then, the estimated motion parameters and geometric distortion field are combined and applied to the slice timing corrected fMRI time series to get the undistorted and re-aligned fMRI data. To deal with involuntary head motion we considered two strategies. First, we used AROMA,^66^ an ICA-based motion correction technique, and then we performed scrubbing using frame-wise displacement > 0.5 mm and root mean squared difference > 0.3 as proposed and validated by Power et al., 2002.^67^ Using the ANTS working field and the fMRI scans in native space all inter- regional functional connectivity for Schaefer atlas is also computed individually for each participant.

#### Processing dMRI Data

The HCP pipeline was used to process our dMRI scans;^68^ briefly, dMRIs were preprocessed by first removing the skull from the b0 image and Topup^65^ was applied to the b0 image to estimate susceptibility distortions. The image set was then passed through an eddy current correction algorithm, the two dMRIs were averaged and a diffusion tensor model fit to each voxel, resulting in a 1st/2nd/3rd eigenvector and eigenvalue map, the fractional anisotropy (FA) map, and mean diffusivity map. Tractography was performed to trace white matter tracks between gray matter regions in a standard Lausanne 156 atlas^69^ + 16 subcortical regions and obtain a structural connectome. Deterministic tractography has been used, followed by global filtering, to minimize false positives; structural connectomes have been constructed as the number of reconstructed fibers between pairs of gray matter regions, normalized by the average regional volume.

### 2.5. Computing TPI components

TPI-li quantifies the effect of local interactions (spatial overlap) between tau and Aβ on tau spread while normalizing for global Aβ/tau burden. Thus, 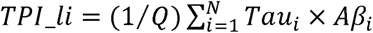, where *Tau_i_* and *Aβ*_i_ Are the Aβ and tau uptake in region-i, 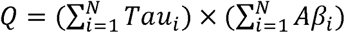 is a normalization factor which controls for the effect of global Aβ and tau deposition at baseline, and *N* is the total number of regions.

Subject-specific Connectomes are utilized in the next three components (TPI-ri, TPI-fc, and TPI-sc). A) Subject-specific functional connectivity matrices are obtained using Schaefer^70^ 200 + 16 subcortical atlas as our functional parcellation. The preprocessed and spatially normalized fMRI data in the MNI template space are used to extract regional time series and subsequently, inter-regional functional connectivity was obtained by calculating the Pearson correlation coefficient (PCC) between all pairs of regions. To eliminate noisy connections, PCCs with absolute values lower than the 30^th^ percentile of the same subjects are eliminated. B) Subject-specific structural connectivity maps are a matrix where each region’s connectivity strength is recorded with all other brain regions in the white matter, based on Lausanne 157^69^ +16 subcortical atlas as our structural parcellation. Thresholding for noisy connections was done by eliminating all tracks with connectivity less than the 25^th^ percentile in the same subject. C) Group-averaged functional and structural connectivity maps were obtained by averaging existing ∼50 young (<40 years) and healthy participants’ subject-specific functional and structural connectivity maps respectively and thresholding them with 30^th^ and 25^th^ percentile.

TPI-ri quantifies the effect of remote interactions between tau and Aβ on the spread of tau. TPI-ri is the weighted average of all possible inter-regional Aβ-tau interactions, normalized by global tau and Aβ deposition. Each inter-regional Aβ-tau interaction was weighted by the subject-specific functional connectivity between the two regions. TPI-ri was based on the reports that tau spread from region-*i* to region-*j* was determined by the amount of tau in region-*i* (*Tau_i_*), the amount of Aβ in region-*j*(*Aβj*), and the functional connectivity between the two regions (*Fij*) which allows the remote interaction to occur. Thus, 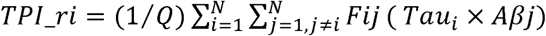, where is the thresholded PCC between regions (described above), *Q* is a normalization factor for global Aβ and tau deposition (defined above), and *N* is the total number of regions. We also explored the calculation of TPI-ri using subject-specific structural connectivity by replacing the *FIJ* in the equation with the structural connectivity matrix, *Sij* Group-averaged TPI-ri computed by group-averaged connectivity maps.

TPI-fc quantifies the effects of functional connectivity on the spread of tau 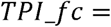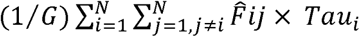, where *F̂ij* is the binarized *Fij*. Essentially, making 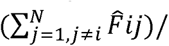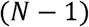 equal to the “hubness” of the region-*i*, reflecting the number of connected regions to region-*i*, as used in prior studies^14,1914^. Dividing by 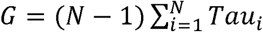, the global tau uptake multiplied by (N − 1) , normalize *TPI_fc* to the [0, 1] range and ensures *TPI_fc* is independent on total tau accumulation at baseline. Essentially, the regions with a greater number of connected regions (higher hubness) resulted in higher *TPI_fc_* In other words, if tau spreads to the regions that are highly connected to the rest of the brain (hub regions) then its progression rate increases substantially. If all regions are connected to all other regions with more than the subject-wise threshold connectivity, *F̂ij* = 1, then *TPI_fc* equal to the maximum value of 1. If there is no functional connectivity between regions, *TPI_fc* becomes zero.

There is ample evidence that structural connectivity can also facilitate spread of tau^15–18,21^ giving rise to the structural connectivity component as 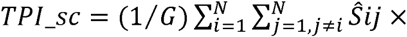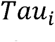, where *S_ij_* is the binarized *S_ij_*, which makes the 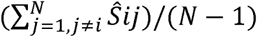 equal to the degree of node-*i* as it was defined in graph theory^71^. The rest of the elements are the same as in *TPI_fc* , and normalization also makes sure *TPI_fc* is within [0,1] range and is independent of the global tau accumulation at baseline.

### 2.6. Statistical Analyses

To evaluate the predictability of the TPI component individually, we perform separate multiple linear regression analyses with individual TPI components as the independent variables of interest, change in global tau as the dependent variable, and control for covariates. To demonstrate the superiority of using subject-specific connectome versus traditional group-averaged connectome, all TPI components were computed using subject-specific connectomes. The significance of the regression coefficient of each TPI component indicated its association with change in global tau over 2.5 years. We repeated the same analysis for each TPI component (except TPI-li which doesn’t rely on any connectome) using group-averaged functional and structural connectivity. We compared the obtained subject-specific TPI components with corresponding group-averaged TPI to assess its stronger association with change in tau. We used the test for two regression coefficients to determine if the slopes were significantly different. We also compared the difference in the explained variance of the global tau change by comparing R^2^ obtained from the two regressions and using an F-test. Once we evaluated all the TPI components, we linearly combined all four TPI components and used linear regression to obtain the best coefficients. This resulted in our final TPI. The association between the final TPI and future tau accumulation was evaluated using the same method we used for each TPI component. In addition, the same method was utilized to demonstrate the superiority of using subject-specific connectome for computing final TPI in comparison to using group-averaged TPI. Sex, age, ICV, race, and ethnicity were considered covariates for all analyses, and a two-sided p-value of less than 0.05 was considered significant.

## 3. Results

### 3.1. Subject-wise and network-wise longitudinal change of tau deposition

In Fig. 1 we demonstrate that change in tau uptake within 2 years can be detected using our tau PET tracer (MK6240) and our advanced processing pipeline. Fig. 1a depicts each subject’s averaged SUVR (Schaefer’s regional parcellation) at the baseline and follow-up visits using spaghetti plots. Each line represents one subject from our cohort. Subjects show a substantial increase in global tau burden (pairwise t=4.32, p<10^-3^; see inserted boxplot in Fig. 1a) as detected by our tau PET scanning and processing pipeline, which highlights the feasibility of detecting changes in global tau accumulation within 2.5 years, as well as marked inter-individual heterogeneity.

**Fig 1.**
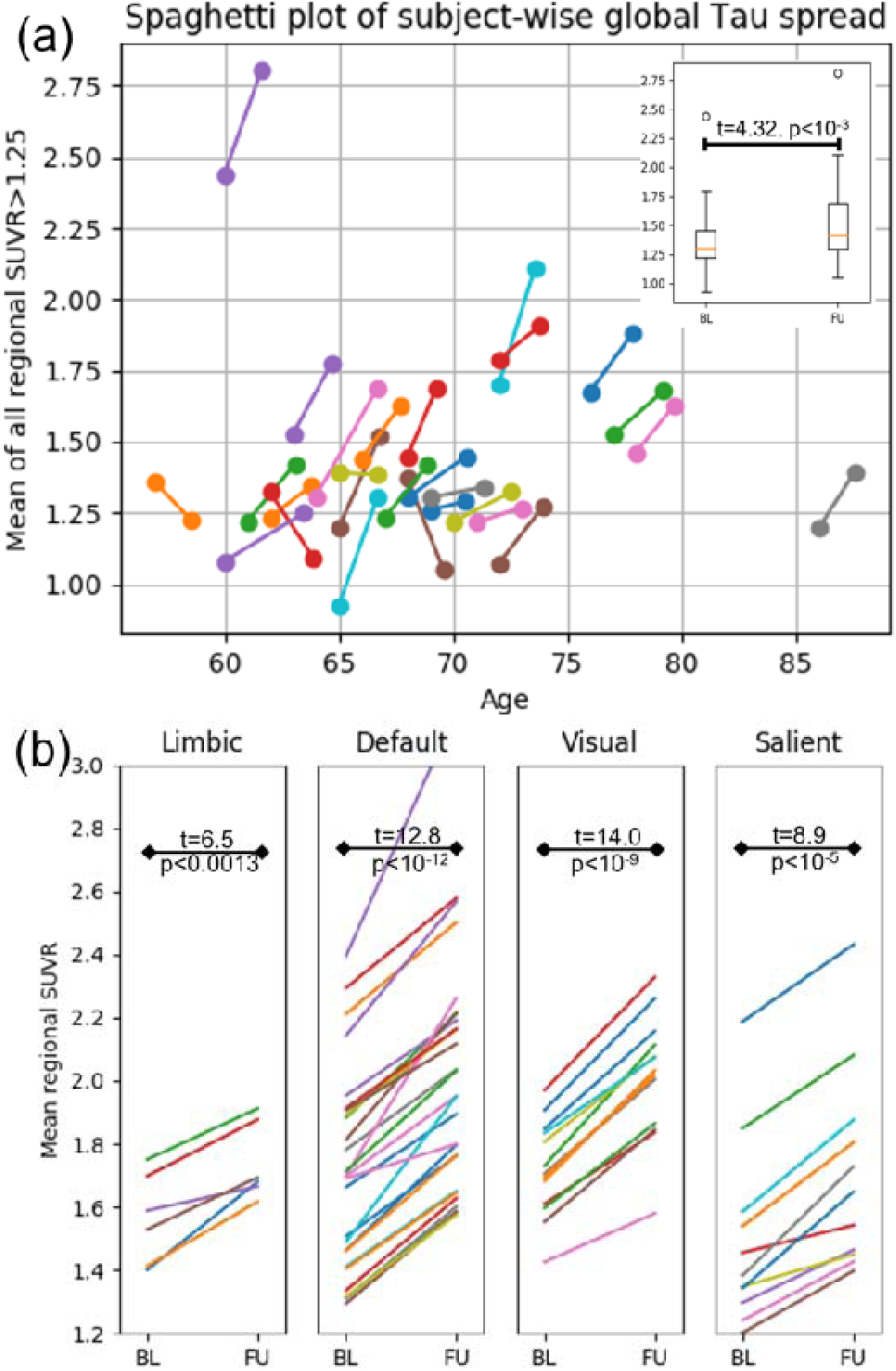
Spaghetti plots illustrating significant tau SUVR increase from baseline (BL) to the 2-year follow-up (FU). a) Subject-wise changes in the global tau SUVR (mean of all regions) in terms of the subject age at BU and FU (each line is one subject). b) Subject-wise changes in the tau SUVR by the network (using Schaefer’s parcellation) are depicted in different plots to highlight the heterogeneity of longitudinal tau change (each line represents one region). Three subjects with marginal global uptake (SUVR<1.4) at baseline showed a decrease in their global tau uptake after 2.5 years. Visual assessment of these tau PET scans revealed significant spill-in artifacts from off-target binding in the skull/meninges during the baseline scan, which decreased on the follow-up.

Next, we aim to show the regional heterogeneity and vulnerabilities in tau accumulation over two years. Fig. 1b illustrates the averaged regional tau SUVR at baseline and follow-up using spaghetti plots. Each line represents the averaged SUVR across all subjects for one region from Schaefer’s atlas. Regions within 4 different networks (Limbic, Default, visual, salient) are depicted in different spaghetti plots. Again, the pair-wise student t-test shows a significant increase in the tau uptake of all 4 networks (t>6.5, p<0.0013). While all regions of the 4 networks show an increase in average tau SUVR, the rate of tau accumulation is extremely heterogeneous across regions. While some regions show a substantially high rate of tau accumulation (ΔSUVR>0.55 per two years), other regions, even with high baseline accumulation, show a very small rate of change (ΔSUVR<0.05 per two years), highlighting regional vulnerability at baseline and their heterogeneous rate of change. Altogether, these results show that change in tau accumulation can be detected during a two-year follow-up period with marked inter-individual and inter-regional heterogeneity, which may be dependent on network connectivity, that we aim to detect using our different TPI components.

### 3.2. The remote interaction component of TPI predicts global longitudinal change of tau deposition

In Fig. 2, TPI-ri is significantly associated with the future tau accumulation (r=0.51, p=0.01) and it accounts for more than 26% of the variance in future tau change. This is also true for the TPI-ri that are computed with group-averaged functional connectivity map (r=0.47, p=0.03); however, the TPI-ri based on subject-specific connectome slightly outperforms the TPI- ri based on group-averaged connectome.

**Fig 2.**
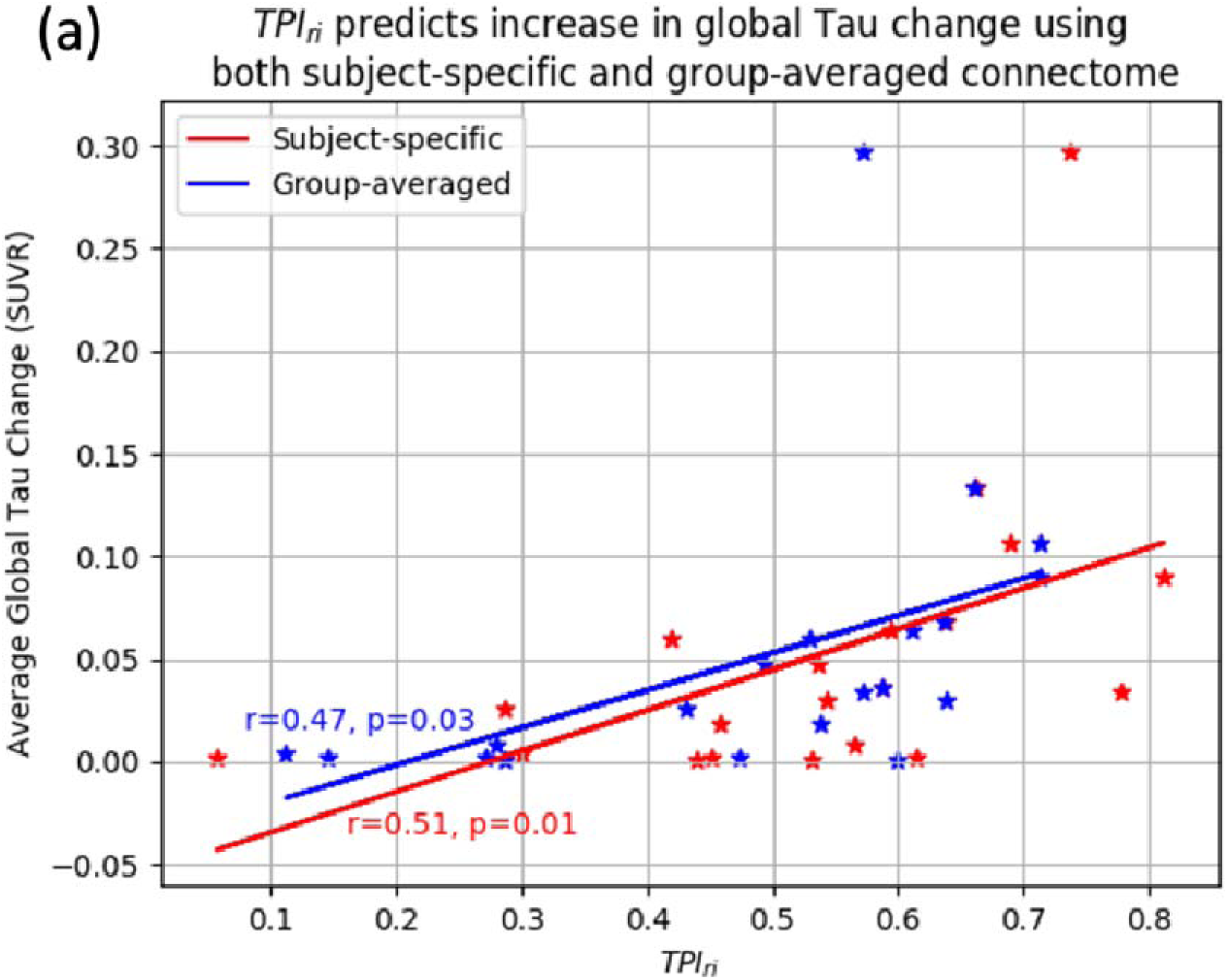
The remote interaction component of our TPI (TPI-ri) predicts global longitudinal change in tau (n=27). The subject-specific connectome (in red) showed slightly increased significance than the group-averaged connectome (in blue).

### 3.3. The local interaction component of TPI (TPI-li) predicts global longitudinal change of tau deposition

Fig. 3 depicts the association between the computed TPI-li and the change in the global tau accumulation. As seen, TPI-li is significantly associated with the change in the global tau accumulation (r=0.49, p=0.02) and it accounts for more than 24% of the variance in the tau change. This high predictability even in this small sample size highlights the robustness of this simple TPI component and is in accordance with our published findings that the multiplication of overlapping Aβ and tau SUVRs is highly associated with neurodegeneration (measured with cortical thickness) independent of total Aβ and tau burden^43^. We have also reported that overlapping Aβ and tau is a more reliable predictor of converting to MCI/AD in HC/MCI participants than the total Aβ and tau pathologies^47^.

**Fig 3.**
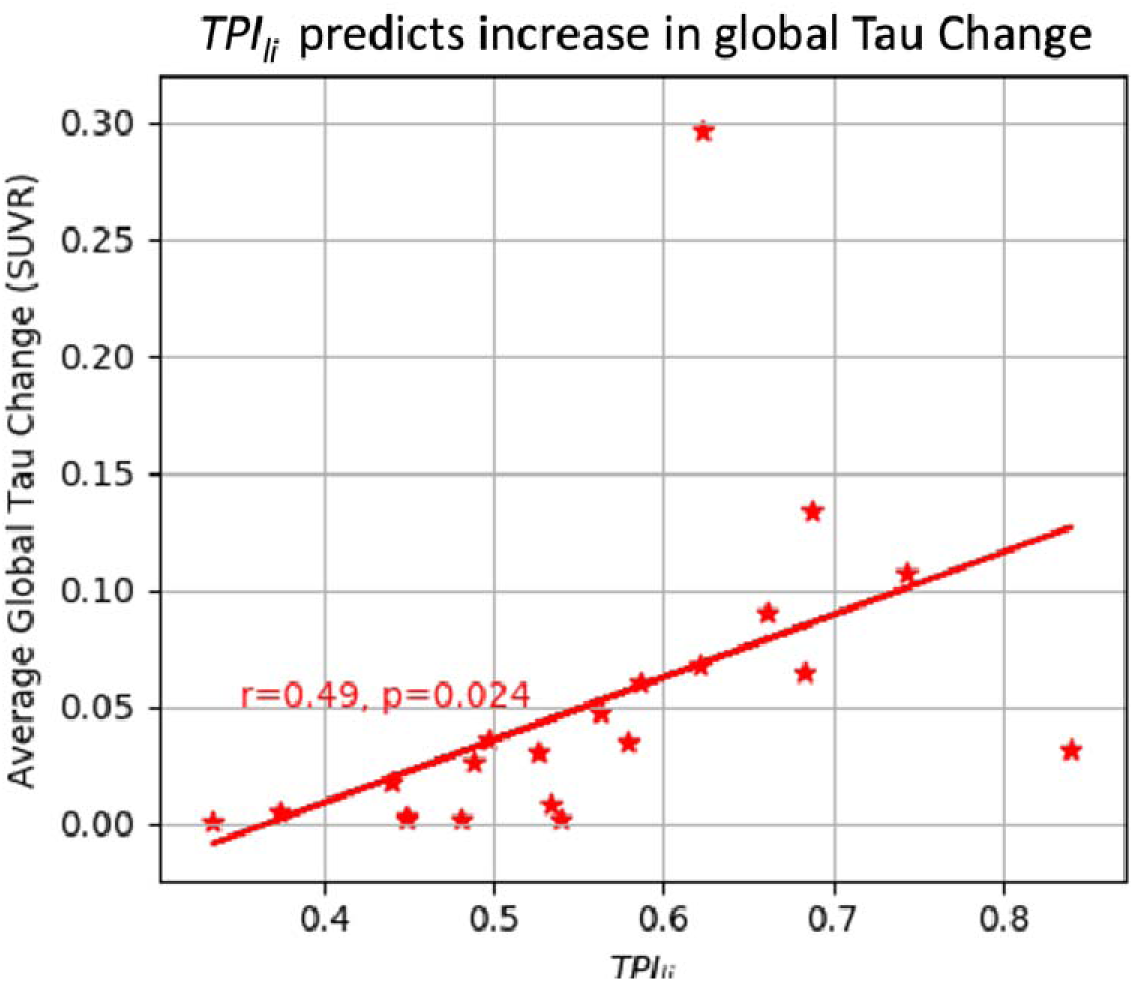
The local interaction component of our TPI (TPI-li) predicts global longitudinal change in tau.

### 3.4. Functional connectivity (TPI-fc) and structural connectivity (TPI-sc) components of TPI predict global longitudinal change of tau deposition

Of the 27 participants, only 21 individuals had rs-fMRI scans acquired with conventional pulse sequences (single 10-min acquisition with TR=2 sec). Thus, we were only able to compute the TPI-fc for these participants. TPI-fc, obtained from the subject-specific connectome, accounts for about 10% of the variance in the tau increase over 2 years, whereas the group-averaged TPI-fc barely accounts for 1% of the variance in tau accumulation increase, and using subject-specific connectome substantially increases the dynamic range of the TPI-fc. These results provide evidence that functional connectivity facilitates the spread of tau, and our proposed TPI-fc can quantify the influence of functional connectivity on the spread of tau in the brain. Furthermore, the TPI-fc computed with subject-specific connectome outperforms the TPI- fc, computed based on group-averaged connectome.

Next, we examined the advantage of using structural connectivity in computing the second component of TPI. Of the 27 participants, only 13 individuals had a dMRI scan with the conventional pulse sequence (single acquisition with 64 directions). Despite the small cohort, we can see the positive trend that TPI-sc predicts an increase in global tau over 2 years (r=0.17 p=0.59). In addition, TPI-sc computed based on subject-specific structural connectivity accounts for more variance in the tau change and has a larger dynamic range than the TPI-sc computed based on group-average structural connectivity.

### 3.5. A combined TPcomponentts predicts longitudinal change of tau deposition

While the two functional and structural connectivity-based TPI components were not significantly associated with future tau accumulation, their inclusion in the final TPI significantly improved the predictability of the final TPI. As seen in Fig. 4, the TPI_final_ obtained from subject-specific connectomes significantly predicts future tau accumulation (r=0.8, p<10^-5^). While the TPI_final_ obtained from group-averaged connectomes also significantly predict future tau accumulation (r=0.58, p<0.007), the subject-specific TPI significantly outperformed the group- averaged TPI_final_ in predicting future tau (DSlope t=2.96, p<0.009) and account for 30% more variance in the future tau accumulation. Achieving such high association using a small sample size in this within-sample validation supports our hypothesis that a single index can in fact accurately predict the future accumulation of tau.

**Fig 4.**
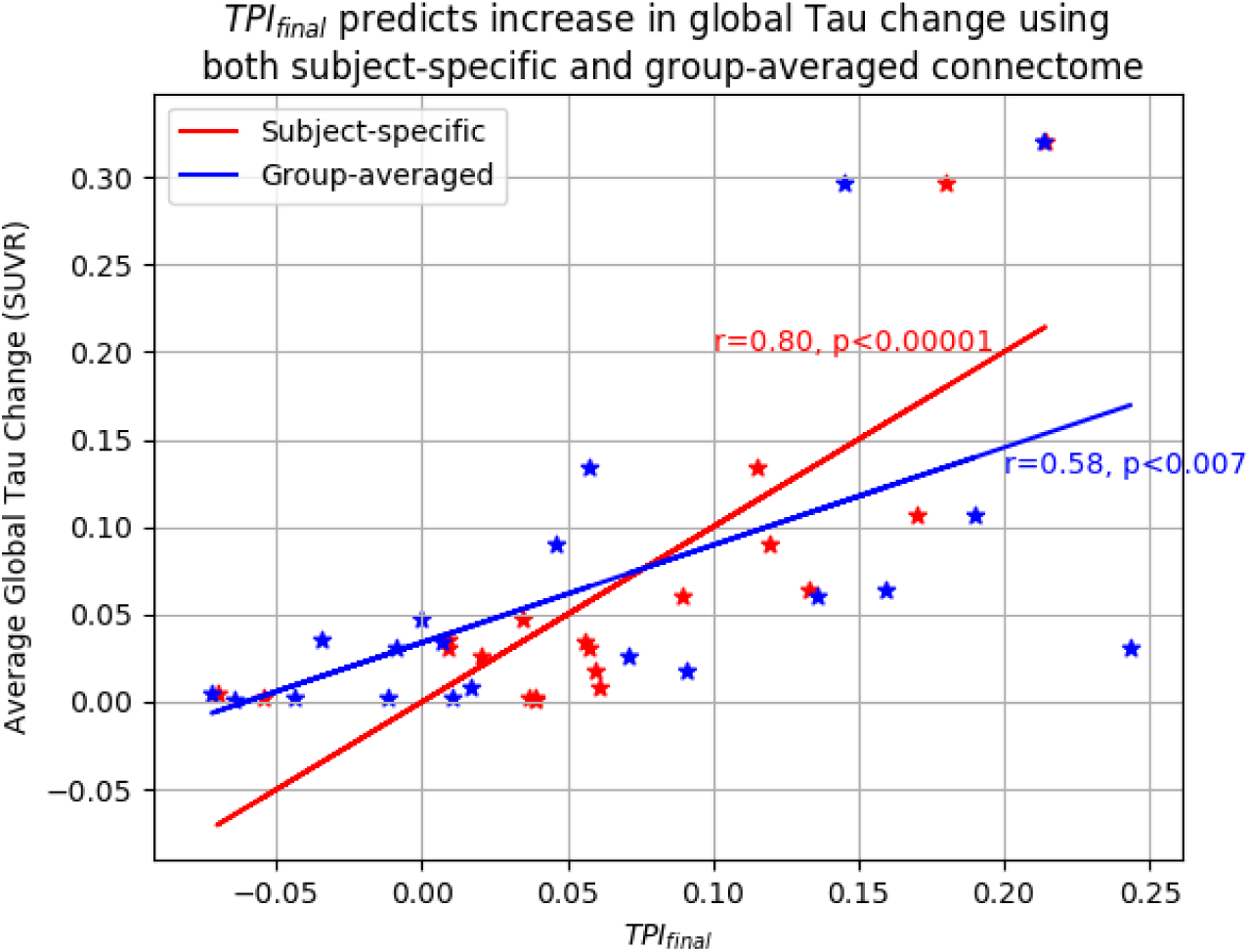
A combined TPI predicts longitudinal change in tau. Using a subject-specific connectome (in red) showed a higher significance than the group-averaged connectome (in blue).

## 4. Discussion

Our novel TPI integrates and advances upon models of tau spread and disease progression proposed by us^42,43,47–49^ and others^14–16,18,72,73^ to generate a unified index encompassing four multimodal components: 1) *TPI-ri* (remote interactions): Aβ/tau interactions in regions where they are functionally or structurally connected but spatially separate,^21,42–46^ 2) *TPI-li* (local interactions): the interactions between co-localized Aβ and tau pathology,^43,44,47^ 3) *TPI-fc:* Subject-specific functional connectivity facilitating tau spread^14–16,48–50^, and 4) *TPI-sc* subject-specific structural connectivity facilitating tau spread.^15–18,21^ Using LASSO fitting and its feature selection capability, for the first time, we build a predictive model that combines Aβ/tau interactions and structural/functional connectivity into a multimodal, multi-component TPI.

There are two previous studies that evaluated the tau progression metrics. First, the network diffusion model (NDM) has shown that relatively simple models of “diffusive” spread recapitulate the spread of tau, both in animal models^74^ and in humans^17,18,75–79^. The core assumption of the NDM is that observed patterns of protein pathology can be largely explained by the trans-neuronal, “prion-like” spread of pathological proteins between regions connected by white matter tracts, which spread along this network along diffusion gradients. Second, the tau hub ratio which receives shows that brain regions that are functionally connected to the regions (hubs) are more influential in spreading the tau than less connected regions (non-hub regions)^14,19^. Using this concept, Frontzkowski et al., 2022 defined tau as the hub ratio to quantify the trajectory of tau spread.^14^ This measure is similar to our TPI-fc except 1) it is computed based on group-averaged functional connectivity, 2) only positive correlations were considered (we consider both strong positive and negative connections), 3) the tau hub ratio is normalized to [-1,1] range whereas our TPI-fc is normalized to [0,1]. Applying these modifications we computed the tau hub ratio using a group-averaged functional connectivity matrix for our comparison and showed that TPI_fc has a better association with future tau accumulation, even though none of the tau hub ratios not TPI_fc predicted future tau accumulation significantly.

Our recent study provided evidence that there are strong associations between Aβ and tau uptake in spatially distinct regions of the brain (remote interactions) during the early stages of AD.^42^ The importance of Aβ/tau remote interaction has been confirmed in animal studies, including demonstration that focal injection of Aβ into a site that already contains tau accelerates both focal tau accumulation and distal tau spread.^80^ The concept of Aβ/tau remote interaction is pivotal to the *amyloid cascade hypothesis,* which proposes that cortical Aβ acts as a gatekeeper for the spread of tau outside the MTL.^81^ However, the network basis of Aβ/tau remote interactions, and how to quantify these interactions in order to predict tau spread and neurodegeneration, is only beginning to be studied. One prior study aimed to use the Aβ and tau burden and structural connectivity to identify the location of the earliest Aβ/tau interaction.^21^ They showed that the inferior temporal gyrus was the region featuring the greatest local amyloid-beta/tau interactions and a connectivity profile well suited to accelerate tau propagation. However, our highly clinically relevant goal is to apply sophisticated image processing/analysis and statistical methods to accurately quantify the effect of these remote interactions on the spread of tau and subsequent disease progression. We hypothesized that the spread of tau from region-1 to region-2 in the brain is determined by tau levels in region-1, Aβ levels in region- 2, and the functional/structural connectivity between the two regions, modulating the Aβ-tau interactions. Our preliminary data indicate that TPI-ri computed based on subject-specific functional connectivity accounts for more than 26% of the variance in the global tau change within 2 years. We have demonstrated that colocalized Aβ and tau within a region (local interaction) predicts future incidence of impairment and dementia^47^, it is also associated with neurodegeneration (measured with cortical thickness), even when controlling for the direct effects of total Aβ and tau burden.^43^ Our preliminary analysis shows that TPI-li can account for more than 24% of the variance in the future accumulation of tau in the brain.

Prior literature investigating the role of connectomes in tau spread used group-averaged connectomes obtained from healthy, young cohorts^14–16,19,21^. However, compelling evidence shows that each individual has a unique brain connectome^31^. Furthermore, connectivity changes with aging^22–26^ and neurodegenerative disease,^27–30^ making connectome templates based on healthy individuals – especially healthy young individuals, as in prior studies – inaccurate. Our use of subject-specific connectomes to account for important inter-individual differences in brain structure and function is novel, justified based on the existing studies, and expected to predict the trajectory of tau spread and cognitive decline/impairment with much greater accuracy on an individual level than is currently possible. Our state-of-the-art image acquisition, processing, and analysis methods allow us to extract accurate and robust individual-based functional and structural connectomes. Our preliminary data show that by using subject-specific connectomes, we are able to account for 30% more variance in the future tau deposition over group-averaged connectomes.

One of the most perplexing issues facing the field of AD investigation is that Aβ and tau proteins can accumulate in individuals who remain asymptomatic for decades or even until the end of life.^2,82^ Understanding why some subjects with evidence of Aβ and tau at autopsy showed no cognitive deficits during life has critical implications for understanding AD pathophysiology.

While we and others have shown that cognitive reserve may partially explain some individuals’ resilience to developing AD,^83–85^ recently network-based hypotheses for the AD pathophysiology (network cascading failure^86,87^, and network degeneration hypothesis^88,89^) are proposed to explain the observed discrepancies between AD pathologies and symptoms. Thus, in this proposal, we propose the novel idea that subject-specific functional and structural brain connectomes and interactions between regional Aβ and tau may play crucial roles in facilitating the spread of tau pathology and subsequent neurodegeneration and cognitive decline. Our proposal builds upon our recent work demonstrating that Aβ and tau interactions are associated with neurodegeneration beyond the direct effects of the total burden of Aβ and tau.^43^ Furthermore, we propose that each individual’s brain functional and structural connectivity not only influences the inter-regional spread of tau but also acts to facilitate the remote interactions between regions containing Aβ and tau^42^. Understanding spatial and network aspects of the contributions of Aβ (A) and tau (T) to neurodegeneration (N) represents a critical refinement to the existing framework that can optimize its explanatory and predictive value in AD research and clinical care in individual patients.^90^

Although our understanding of the molecular basis of AD has advanced significantly in recent years, forming the basis for the 2018 NIA-AA (National Institute of Aging and Alzheimer’s Association) Research Framework^91^, our ability to predict disease progression remains elusive.^92^ We and others have demonstrated marked heterogeneity in longitudinal atrophy patterns and cognitive decline despite the presence of both Aβ and tau pathology on PET imaging^13,47,73,92–95^. On an individual patient level, this unpredictable heterogeneity makes it difficult to convey prognosis to families and caregivers. In clinical trials, this heterogeneity within study populations affects trial outcomes, necessitating larger sample sizes and weakening the perceived efficacy of therapeutic interventions^92^. While tau imaging has been shown to improve prediction of clinical/cognitive trajectories,^5–8^ it remains suboptimal, predicting at best 53% of the variance in cognitive decline^73,95^.

Our main limitations are first, our sample size is rather limited; However, recruiting only non-demented and asymptomatic preclinical AD patients with confirmed early AD imaging pathologies makes our samples substantially less heterogeneous than conventional sampling from the community. The reduction in the variability of our sample size resulted in much higher statistical power and eventually required fewer samples for this study. As it is evident from our highly significant findings. With conventional definition, our sample size could be considered 650; However, including subjects with no tau and Aβ accumulation or subjects with widespread accumulation of tau and Aβ at baseline only diminishes our ability to effectively evaluate our proposed TPI measurement. Having said that, further evaluation of our proposed TPI with a much larger sample size increases the confidence about the predictability of the TPI. Second, due to the small sample size, we were not able to evaluate or propose TPI using conventional methods of within-sample validations. Instead, we used the LASSO model to fit the TPI and performed a within-sample validation by investigating the association of the fitted TPI to future tau accumulation. However, in the future utilizing 20-fold cross-validation with 2000 iterations with different but random selections of the training and testing samples as well as out-of-sample validation using a completely different cohort is warranted to have full confidence in the effectiveness of our proposed TPI for predicting future tau accumulation. Third, using the LASSO model to combine the TPI components into a TPI final may best predict change in global tau but not the clinical outcomes (cortical thinning, cognitive decline, and conversion to MCI/AD). An alternative approach would be to use principal component analysis, which could be tested against changes in global tau and clinical outcomes in the future. Finally, we do not expect to see a substantial number of participants starting monoclonal Aβ before the end of their participation period in this study, since we are enrolling preclinical AD subjects.

## Data Availability

The data for this project are confidential but may be obtained with Data Use Agreements with the Weill Cornell Medicine and Columbia University Irving Medical Center. It can take some weeks to negotiate data use agreements and gain access to the data. The author will assist with any reasonable replication attempts for the following publication.

## Acknowledgments

Our sincere appreciation goes out to all those who played a part in enabling this neuroimaging study to take place. We would first like to express our gratitude to the participants who voluntarily underwent the scanning process, without whom this study would not have been possible. We are grateful for their cooperation, patience, and time invested in the scanning procedures. Finally, we would like to acknowledge all the staff who provided support during the neuroimaging scans. Their professionalism and expertise were invaluable in ensuring the accuracy and reliability of the data.

## Consent Statement

All human participants provided informed consent.

## References

1. Association, A. Alzheimer’s Association Report: 2020 Alzheimer’s disease facts and figures. Alzheimers Dement. J. Alzheimers Assoc. 16, 391–460 (2020).

2. Jagust, W. Imaging the evolution and pathophysiology of Alzheimer’s disease. Nat. Rev. Neurosci. 19, 687–700 (2018).

3. Lloret, A. et al. When does Alzheimer′s disease really start? The role of biomarkers. International Journal of Molecular Sciences vol. 20 (2019).

4. M.D. David S. Knopman, M. D. C. R. J. J. et al. Update on hypothetical model of Alzheimer’s disease biomarkers. 12, 207–216 (2014).

5. Brier, M. R. et al. Tau and Aβ imaging, CSF measures, and cognition in Alzheimer’s disease. Sci. Transl. Med. 8, (2016).

6. Gordon, B. A. et al. Cross-sectional and longitudinal atrophy is preferentially associated with tau rather than amyloid β positron emission tomography pathology. *Alzheimer’s Dement. Diagnosis*, Assess. Dis. Monit. 10, 245 (2018).

7. Bucci, M., Chiotis, K. & Nordberg, A. Alzheimer’s disease profiled by fluid and imaging markers: tau PET best predicts cognitive decline. Mol. Psychiatry 2021 2610 26, 5888–5898 (2021).

8. Biel, D. et al. Tau-PET and in vivo Braak-staging as prognostic markers of future cognitive decline in cognitively normal to demented individuals. Alzheimer’s. Res. Ther. 13, (2021).

9. Iacono, D. et al. Mild Cognitive Impairment and Asymptomatic Alzheimer Disease Subjects: Equivalent A-Amyloid and Tau Loads With Divergent Cognitive Outcomes. J. Neuropathol. Exp. Neurol. 73, 295–304 (2014).

10. Perez-Nievas, B. G. et al. Dissecting phenotypic traits linked to human resilience to Alzheimer’s pathology. Brain 136, 2510–2526 (2013).

11. Lowe, V. J. et al. Elevated medial temporal lobe and pervasive brain tau-PET signal in normal participants. *Alzheimer’s Dement. Diagnosis*, Assess. Dis. Monit. 10, 210–216 (2018).

12. Jack, C. R. et al. Associations of Amyloid, Tau, and Neurodegeneration Biomarker Profiles with Rates of Memory Decline among Individuals Without Dementia. JAMA - Journal of the American Medical Association vol. 321 2316–2325 (2019).

13. Strikwerda-Brown, C. et al. Association of Elevated Amyloid and Tau Positron Emission Tomography Signal with Near-Term Development of Alzheimer Disease Symptoms in Older Adults Without Cognitive Impairment. JAMA Neurol. 79, 975–985 (2022).

14. Frontzkowski, L. et al. Earlier Alzheimer’s disease onset is associated with tau pathology in brain hub regions and facilitated tau spreading. Nat. Commun. 13, 1–14 (2022).

15. Vogel, J. W. et al. Spread of pathological tau proteins through communicating neurons in human Alzheimer’s disease. Nat. Commun. 11, 2612 (2020).

16. Therriault, J. et al. Intrinsic connectivity of the human brain provides scaffold for tau aggregation in clinical variants of Alzheimer’s disease. Sci. Transl. Med. 14, eabc8693 (2022).

17. Raj, A. et al. Network Diffusion Model of Progression Predicts Longitudinal Patterns of Atrophy and Metabolism in Alzheimer’s Disease. Cell Rep. 10, 359–369 (2015).

18. Raj, A., Kuceyeski, A. & Weiner, M. A Network Diffusion Model of Disease Progression in Dementia. Neuron 73, 1204–1215 (2012).

19. Franzmeier, N. et al. Patient-centered connectivity-based prediction of tau pathology spread in Alzheimer’s disease. Sci. Adv. 6, (2020).

20. Wang, Y. et al. The release and trans-synaptic transmission of Tau via exosomes. Mol. Neurodegener. 12, (2017).

21. Lee, W. J. et al. Regional Aβ-tau interactions promote onset and acceleration of Alzheimer’s disease tau spreading. Neuron 110, 1932–1943.e5 (2022).

22. Razlighi, Q. R. et al. Unilateral disruptions in the default network with aging in native space. Brain Behav. 4, (2014).

23. Varangis, E., Habeck, C. G., Razlighi, Q. R. & Stern, Y. The Effect of Aging on Resting State Connectivity of Predefined Networks in the Brain. Front. Aging Neurosci. 11, (2019).

24. Tomasi, D. & Volkow, N. D. Aging and functional brain networks. Mol. Psychiatry 17, 549–558 (2011).

25. Liem, F., Geerligs, L., Damoiseaux, J. S. & Margulies, D. S. Functional connectivity in aging. *Handb*. Psychol. Aging 37–51 (2021) doi:10.1016/B978-0-12-816094-7.00010-6.

26. Andrews-Hanna, J. R. et al. Disruption of large-scale brain systems in advanced aging. Neuron 56, 924–35 (2007).

27. Sheline, Y. I. & Raichle, M. E. Resting state functional connectivity in preclinical Alzheimer’s disease. Biol. Psychiatry 74, 340–347 (2013).

28. Rombouts, S. A. R. B., Barkhof, F., Goekoop, R., Stam, C. J. & Scheltens, P. Altered resting state networks in mild cognitive impairment and mild Alzheimer’s disease: An fMRI study. Hum. Brain Mapp. 26, 231–239 (2005).

29. Binnewijzend, M. A. A. et al. Resting-state fMRI changes in Alzheimer’s disease and mild cognitive impairment. Neurobiol. Aging 33, 2018–2028 (2012).

30. Greicius, M. D., Srivastava, G., Reiss, A. L. & Menon, V. Default-mode network activity distinguishes Alzheimer’s disease from healthy aging: evidence from functional MRI. Proc Natl Acad Sci U S A 101, 4637–4642 (2004).

31. Shen, X. et al. Functional connectome fingerprinting: identifying individuals using patterns of brain connectivity. Nat. Neurosci. 18, 1–11 (2015).

32. Feinberg, D. A. & Setsompop, K. Ultra-fast MRI of the human brain with simultaneous multi-slice imaging. J. Magn. Reson. 229, 90–100 (2013).

33. Preibisch, C., Castrillón G., J. G., Bührer, M. & Riedl, V. Evaluation of Multiband EPI Acquisitions for Resting State fMRI. PLoS One 10, (2015).

34. Parker, D. & Razlighi, Q. Attenuation of motion artifacts in fMRI using discrete reconstruction of irregular fMRI trajectories (DRIFT). Magn. Reson. Med. 1–14 (2021) doi:10.1101/714436.

35. Parker, D., Liu, X. & Razlighi, Q. R. Optimal slice timing correction and its interaction with fMRI parameters and artifacts. Med. Image Anal. 35, 434–445 (2016).

36. He, H. & Razlighi, Q. R. Landmark-guided region-based spatial normalization for functional magnetic resonance imaging. Hum. Brain Mapp. 43, 3524 (2022).

37. Mejia, A. F., Nebel, M. B., Wang, Y., Caffo, B. S. & Guo, Y. Template Independent Component Analysis: Targeted and Reliable Estimation of Subject-level Brain Networks using Big Data Population Priors. J. Am. Stat. Assoc. 115, 1151 (2020).

38. Smith, S. M. et al. Network modelling methods for FMRI. Neuroimage 54, 875–91 (2011).

39. Glasser, M. F. et al. A multi-modal parcellation of human cerebral cortex. Nature 1–11 (2016) doi:10.1038/nature18933.

40. Beckmann, C. F. & Smith, S. M. Probabilistic Independent Component Analysis for Functional Magnetic Resonance Imaging. IEEE Trans. Med. Imaging 23, 137–152 (2004).

41. Damoiseaux, J. S. et al. Consistent resting-state networks across healthy subjects. Proc. Natl. Acad. Sci. U. S. A. 103, 13848–13853 (2006).

42. Hojjati, S. H. et al. Disentangling the distal association between β-Amyloid and tau pathology at varying stages of tau deposition. medRxiv 2023.03.31.23288013 (2023) doi:10.1101/2023.03.31.23288013.

43. Hani Hojjati, S., et al. Distinct and joint effects of low and high levels of Aβ and tau deposition on cortical thickness. NeuroImage Clin. 38, (2023).

44. Iaccarino, L. et al. Local and distant relationships between amyloid, tau and neurodegeneration in Alzheimer’s Disease. NeuroImage Clin. 17, 452–464 (2018).

45. Busche, M. A. & Hyman, B. T. Synergy between amyloid-β and tau in Alzheimer’s disease. Nat. Neurosci. 23, 1183–1193 (2020).

46. Pascoal, T. A. et al. Synergistic interaction between amyloid and tau predicts the progression to dementia. Alzheimer’s Dement. 13, 644–653 (2017).

47. Hojjati, S. H., Feiz, F., Ozoria, S. & Razlighi, Q. R. Topographical Overlapping of the Amyloid-β and Tau Pathologies in the Default Mode Network Predicts Alzheimer’s Disease with Higher Specificity. J. Alzheimer’s Dis. 83, 407–421 (2021).

48. Nayak, S., et al. Functional Connectivity of Entorhinal Cortex Predicts Tau in the Elderly. in Alzheimer Association International Conference (AAIC) (2023).

49. Razlighi, Q. R., Chiang, G., Nayak, S. & Hojjati, H. Regions with higher functional connectivity to the medial temporal lobe have higher tau accumulation even after controlling for their Euclidean distance. in Socity for Neuroscience (2023).

50. Franzmeier, N. et al. Patient-centered connectivity-based prediction of tau pathology spread in Alzheimer’s disease. Sci. Adv. 6, (2020).

51. Albert, M. S. et al. The diagnosis of mild cognitive impairment due to Alzheimer’s disease: Recommendations from the National Institute on Aging-Alzheimer’s Association workgroups on diagnostic guidelines for Alzheimer’s disease. Alzheimer’s Dement. 7, 270–279 (2011).

52. Fischl, B. et al. Whole brain segmentation: automated labeling of neuroanatomical structures in the human brain. Neuron 33, 341–355 (2002).

53. Fischl, B. et al. Automatically parcellating the human cerebral cortex. Cereb. Cortex 14, 11–22 (2004).

54. Desikan, R. S. et al. An automated labeling system for subdividing the human cerebral cortex on MRI scans into gyral based regions of interest. Neuroimage 31, 968–80 (2006).

55. Fischl, B. & Dale, a M. Measuring the thickness of the human cerebral cortex from magnetic resonance images. Proc. Natl. Acad. Sci. U. S. A. 97, 11050–11055 (2000).

56. Ardekani, B. A., Braun, M., Hutton, B. F., Kanno, I. & Iida, H. A fully automatic multimodality image registration algorithm. J. Comput. Assist. Tomogr. 19, 615–623 (1995).

57. Reuter, M., Schmansky, N. J., Rosas, H. D. & Fischl, B. Within-subject template estimation for unbiased longitudinal image analysis. Neuroimage 61, 1402–1418 (2012).

58. Tahmi, M., Bou-Zeid, W. & Razlighi, Q. R. A fully automatic technique for precise localization and quantification of amyloid-b PET scans. J. Nucl. Med. 60, (2019).

59. Oh, H., Steffener, J., Razlighi, Q. R., Habeck, C. & Stern, Y. β-Amyloid Deposition Is Associated with Decreased Right Prefrontal Activation during Task Switching among Cognitively Normal Elderly. J. Neurosci. 36, 1962–70 (2016).

60. Oh, H. et al. AB-related hyperactivation in frontoparietal control regions in cognitively normal elderly. Neurobiol. Aging 36, 3247–3254 (2015).

61. Brickman, A. M. et al. Cerebral autoregulation, beta amyloid, and white matter hyperintensities are interrelated. Neurosci. Lett. 592, 54–58 (2015).

62. Gu, Y. et al. Brain amyloid deposition and longitudinal cognitive decline in nondemented older subjects: Results from a multi-ethnic population. PLoS One 10, 1–14 (2015).

63. Parker, D., Gerraty, R. T. & Razlighi, Q. R. Optimal signal recovery from interleaved FMRI data. in Proceedings - International Symposium on Biomedical Imaging vols 2015-July (2015).

64. Parker, D., Rotival, G., Laine, A. & Razlighi, Q. R. Retrospective detection of interleaved slice acquisition parameters from fMRI data. in 2014 IEEE 11th International Symposium on Biomedical Imaging, ISBI 2014 (2014).

65. Andersson, J. L. R., Skare, S. & Ashburner, J. How to correct susceptibility distortions in spin-echo echo-planar images: Application to diffusion tensor imaging. Neuroimage 20, 870–888 (2003).

66. Pruim, R. H. R. et al. ICA-AROMA: A robust ICA-based strategy for removing motion artifacts from fMRI data. Neuroimage 112, 267–277 (2015).

67. Power, J. D., Barnes, K. a, Snyder, A. Z., Schlaggar, B. L. & Petersen, S. E. Spurious but systematic correlations in functional connectivity MRI networks arise from subject motion. Neuroimage 59, 2142–54 (2012).

68. Glasser, M. F. et al. The minimal preprocessing pipelines for the Human Connectome Project. Neuroimage 80, 105–124 (2013).

69. Cammoun, L. et al. Mapping the human connectome at multiple scales with diffusion spectrum MRI. J. Neurosci. Methods 203, 386–397 (2012).

70. Schaefer, A. et al. Local-Global Parcellation of the Human Cerebral Cortex from Intrinsic Functional Connectivity MRI. Cereb. Cortex 28, 3095–3114 (2018).

71. Diestel, R. Graph Theory(5th Edition). Springer (2017).

72. Strikwerda-Brown, C. et al. Association of Elevated Amyloid and Tau Positron Emission Tomography Signal With Near-Term Development of Alzheimer Disease Symptoms in Older Adults Without Cognitive Impairment. JAMA Neurol. 79, 975–985 (2022).

73. Vogel, J. W. et al. Four distinct trajectories of tau deposition identified in Alzheimer’s disease. Nat. Med. (2021) doi:10.1038/s41591-021-01309-6.

74. Mezias, C. & Raj, A. Analysis of Amyloid-β pathology spread in mouse models suggests spread is driven by spatial proximity, not connectivity. Front. Neurol. 8, (2017).

75. Acosta, D., Powell, F., Zhao, Y. & Raj, A. Regional vulnerability in Alzheimer’s disease: The role of cell-autonomous and transneuronal processes. Alzheimer’s Dement. 14, (2018).

76. Torok, J., Maia, P. D., Powell, F., Pandya, S. & Raj, A. A method for inferring regional origins of neurodegeneration. Brain 141, (2018).

77. Freeze, B., Maia, P., Pandya, S. & Raj, A. Network mediation of pathology pattern in sporadic Creutzfeldt-Jakob disease. Brain Commun. 2, (2020).

78. Pandya, S., Mezias, C. & Raj, A. Predictive model of spread of progressive supranuclear palsy using directional network diffusion. Front. Neurol. 8, (2017).

79. Pandya, S. et al. Modeling seeding and neuroanatomic spread of pathology in amyotrophic lateral sclerosis. Neuroimage 251, (2022).

80. Lasagna-Reeves, C. A. et al. Alzheimer brain-derived tau oligomers propagate pathology from endogenous tau. Sci. Rep. 2, (2012).

81. Hardy, J. A. & Higgins, G. A. Alzheimer’s disease: the amyloid cascade hypothesis. Science 256, 184–185 (1992).

82. Dickerson, B. C. & Sperling, R. A. Neuroimaging biomarkers for clinical trials of disease-modifying therapies in Alzheimer’s disease. NeuroRx 2, 348–360 (2005).

83. Oh, H., Razlighi, Q. R. & Stern, Y. Multiple pathways of reserve simultaneously present in cognitively normal older adults. Neurology 90, e197 LP-e205 (2018).

84. Habeck, C. et al. Cognitive Reserve and Brain Maintenance: Orthogonal Concepts in Theory and Practice. Cereb. Cortex 1–8 (2016) doi:10.1093/cercor/bhw208.

85. Stern, Y., Gazes, Y., Razlighi, Q., Steffener, J. & Habeck, C. A task-invariant cognitive reserve network. Neuroimage 178, (2018).

86. Jones, D. T. et al. Tau, amyloid, and cascading network failure across the Alzheimer’s disease spectrum. Cortex 97, 143–159 (2017).

87. Jones, D. T. et al. Cascading network failure across the Alzheimer’s disease spectrum. Brain 139, 547–562 (2016).

88. Alexander Drzezga. The Network Degeneration Hypothesis: Spread of Neurodegenerative Patterns Along Neuronal Brain Networks. J. Nucl. Med. 59, 1645– 1648 (2018).

89. Hoenig, M. C. et al. Networks of tau distribution in Alzheimer’s disease. Brain 141, 568–581 (2018).

90. Ricciarelli, R. & Fedele, E. The Amyloid Cascade Hypothesis in Alzheimer’s Disease: It’s Time to Change Our Mind. Curr. Neuropharmacol. 15, (2017).

91. Jack, C. R. et al. NIA-AA Research Framework: Toward a biological definition of Alzheimer’s disease. Alzheimer’s Dement. 14, 535–562 (2018).

92. Duara, R. & Barker, W. Heterogeneity in Alzheimer’s Disease Diagnosis and Progression Rates: Implications for Therapeutic Trials. Neurotherapeutics 19, 8 (2022).

93. Simon, S. S. et al. In vivo tau is associated with change in memory and processing speed, but not reasoning, in cognitively unimpaired older adults. Neurobiol. Aging 133, 28–38 (2024).

94. Kreisl, W. C. et al. Patterns of tau pathology identified with 18F-MK-6240 PET imaging. Alzheimer’s Dement. 18, 272–282 (2022).

95. Ossenkoppele, R. et al. Accuracy of Tau Positron Emission Tomography as a Prognostic Marker in Preclinical and Prodromal Alzheimer Disease: A Head-to-Head Comparison Against Amyloid Positron Emission Tomography and Magnetic Resonance Imaging. JAMA Neurol. 78, 961–971 (2021).

